# Automated biometry for assessing cephalopelvic disproportion in 3D 0.55T fetal MRI at term

**DOI:** 10.1101/2025.08.18.25333914

**Authors:** Alena U. Uus, Simi Bansal, Yagmur Gerek, Hadi Waheed, Sara Neves Silva, Jordina Aviles Verdera, Vanessa Kyriakopoulou, Lia Betti, Shireen Jaufuraully, Joseph V. Hajnal, Dimitris Siasakos, Anna David, Manju Chandiramani, Jana Hutter, Lisa Story, Mary A. Rutherford

## Abstract

Fetal MRI offers detailed three-dimensional visualisation of both fetal and maternal pelvic anatomy, allowing for assessment of the risk of cephalopelvic disproportion and obstructed labour. However, conventional measurements of fetal and pelvic proportions and their relative positioning are typically performed manually in 2D, making them time-consuming, subject to inter-observer variability, and rarely integrated into routine clinical workflows.

In this work, we present the first fully automated pipeline for pelvic and fetal head biometry in T2-weighted fetal MRI at late gestation. The method employs deep learning-based localisation of anatomical landmarks in 3D reconstructed MRI images, followed by computation of 12 standard linear and circumference measurements commonly used in the assessment of cephalopelvic disproportion. Landmark detection is based on 3D UNet models within MONAI framework, trained on 57 semi-manually annotated datasets.

The full pipeline is quantitatively validated on 10 test cases. Furthermore, we demonstrate its clinical feasibility and relevance by applying it to 206 fetal MRI scans (36–40 weeks’ gestation) from the MiBirth study, which investigates prediction of mode of delivery using low field MRI.

## 1 Introduction

Cephalopelvic disproportion (CPD) and labour dystocia are common indications for emergency caesarean delivery. These complications are linked to the spatial relationship between the fetal head and maternal pelvis.

Recent advances in fetal MRI, allow acquisition of high-quality 3D images of the uterus and pelvis at late gestation. This enables direct volumetric assessment of clinically relevant fetal and maternal anatomical features, including fetal head dimensions and pelvic diameters [5]. Although these parameters have demonstrated utility in assessing CPD risks [11,7,1], they are not employed in routine clinical practice due to the complexity and time requirements of manual annotations, as well as the lack of clinical validation studies and formalised normative ranges. Furthermore, manual measurements are traditionally performed in 2D slices and are affected by inter-observer variability and limited reproducibility.

While deep learning has shown promising results for automated fetal brain biometry for 3D MRI [10,9], no existing methods address the automation of maternal pelvic measurements or estimation of combined fetopelvic indices in fetal MRI. This limits the clinical utility of fetal MRI for labour risk stratification.

## Contributions

In this work, we present the first automated solution for assessing measurements associated with cephalopelvic disproportion in late gestation low field fetal MRI. The method performs fully automated biometry of the maternal pelvis and fetal head according to standard clinical measurement protocols, followed by computation of the cephalopelvic index. Measurements are derived from deep learning-based localisation of 12 pelvic and 6 fetal head landmarks in 3D reconstructed T2-weighted 0.55T MRI images of the pelvis and uterus ROI. The landmark detection networks are based on 3D UNet architectures and trained on 57 datasets with semi-manually annotated ground-truth labels. The pipeline is quantitatively evaluated on 10 independent cases and subsequently applied to assess cephalopelvic parameters in 206 late gestation 0.55T MRI datasets.

## 2 Methods

### 2.1 Cohort, acquisition and pre-processing

This study employs fetal MRI data acquired at St.Thomas’ Hospital, London, UK under the “MiBirth: MRI imaging at term for prediction of the mode of birth” study (REC: 23/LO/0685) that specifically focuses on prediction of the mode of delivery using low field imaging. It includes 206 normal control datasets from 36-401 weeks gestational age (GA) range. All experiments were performed in accordance with relevant guidelines and regulations. Informed written consent was obtained from all participants.

The fetal datasets were acquired on a 0.55T clinical MRI scanner (MAGNETOM Free.Max, Siemens Healthcare, Germany) in the supine position, using a 6-element blanket coil and a 9-element spine coil. In each datasets, 8-11 T2w stacks covering the whole uterus, pelvis and fetal head were acquired with an optimised ssTSE sequence [2] TE=105–106 ms, acquisition resolution 1.48×1.48 mm, slice thickness 4.5 mm.

The 3D images of the whole uterus+pelvis ROI were reconstructed to 1.5mm isotropic resolution using in-house automated pipeline based on deformable slice-to-volume reconstruction (DSVR) [12,13] in SVRTK package^9^ with deep learning masking.

The inclusion criteria were: no reported fetal or maternal anomalies, singleton pregnancy, cephalic presentation and sufficient reconstruction image quality with acceptable visibility of the pelvis and fetal head.

### 2.2 Formalisation of cephalopelvic biometry protocol

The proposed protocol for fetopelvic biometry for 3D T2w MRI is based on conventional linear measurements used in assessment of cephalopelvic disproportion, pelvimetry and fetal head biometry [8]. It was formalised based on a series of consensus meetings between clinicians and researchers (MR, SJ, SB, AU) with extensive experience in fetal MRI and obstetrics. The pelvis measurements include 6 distances and 2 circumferences. The fetal head measurements include 3 distances and 1 circumference. The circumferences are computed as *π* (*d*_1_ + *d*_2_)*/*2 (where *d*_1_ and *d*_2_ are diameters). Next, these measurements were defined via landmark-based approach in 3D T2w reconstructed images of the pelvis and uterus ROI. The landmarks were created as 3D sphere labels using ITK-SNAP^10^ and 3DSlicer^11^ software. In order to facilitate localisation of the landmarks, parcellation of the pelvic bones and fetal head ROIs were additionally added to the protocol.

### 2.3 Automated cephalopelvic biometry pipeline

The proposed pipeline for automated cephalopelvic biometry is summarised in Fig. 1. Initially, anatomical landmarks are localised in the 3D T2-weighted DSVR images of the pelvis and uterus ROI using two dedicated 3D segmentation networks - one for fetal landmarks and one for pelvic landmarks. The extracted landmark coordinates are then used to compute linear distances, which are subsequently used to derive circumferential measurements.

**Fig. 1.**
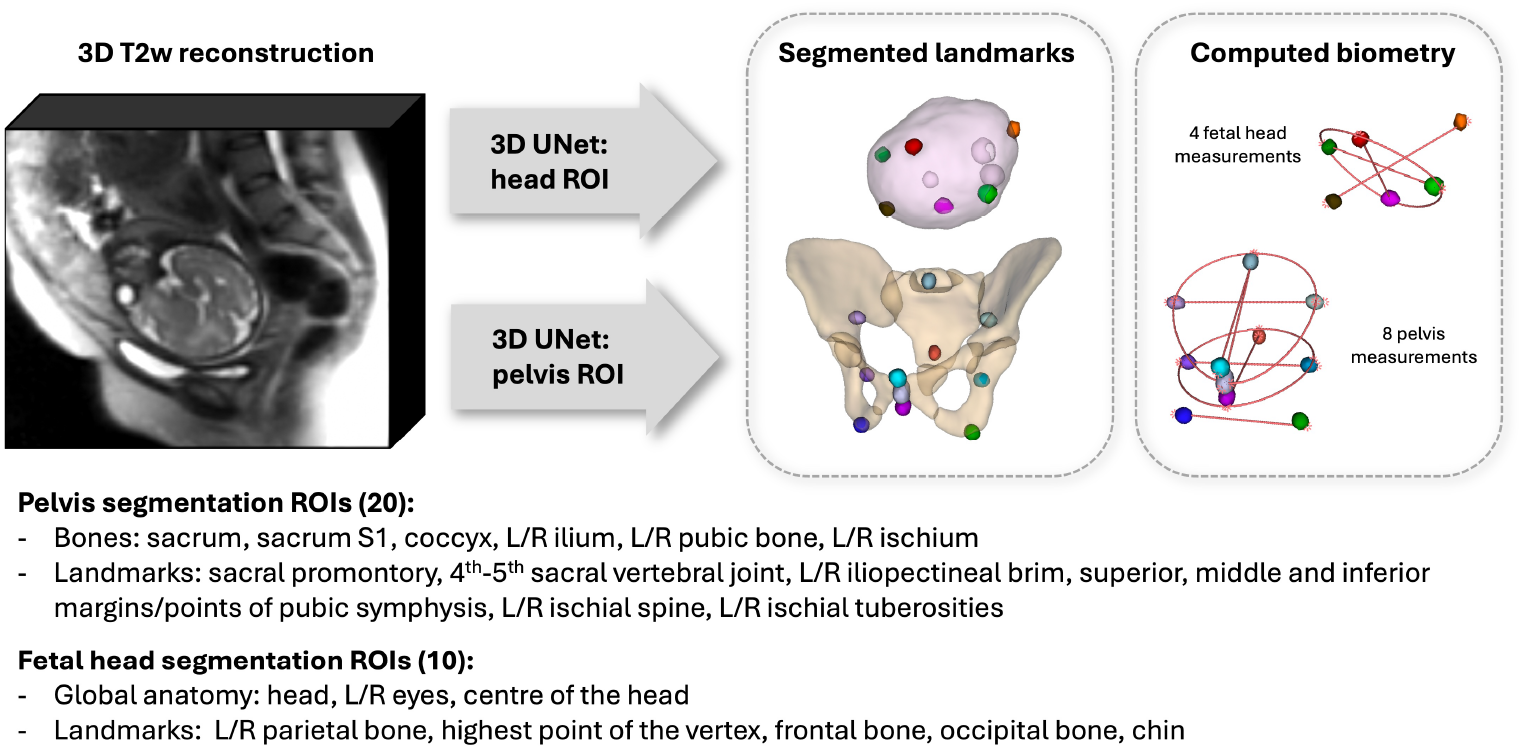
Proposed pipeline for automated cephalopelvic biometry for 3D T2w fetal MRI.

For the segmentation networks, we employed the standard MONAI [3] implementation of the 3D UNet [14], configured with five and four encoder–decoder blocks and output channels of 32, 64, 128, 256, and 512. Both convolution and upsampling used a kernel size of 3, with ReLU activation and a dropout ratio of 0.5. Optimisation was performed using the AdamW optimiser with default *β* parameters, a weight decay of 1 *×* 10^*−*5^, and a linearly decaying learning rate initialised at 1 *×* 10^*−*3^. Data augmentation followed MONAI standard protocol, including affine rotations, contrast adjustment, and bias field simulation. Preprocessing included transformation to a common reference space, cropping to the pelvis and head ROI, intensity normalisation to 0–1 range, and resampling with padding to a uniform 128×128×128 voxel grid.

The training and testing datasets were generated semi-manually using internal in-house deep learning segmentation models to pre-label pelvic bones and the fetal head, followed by registration-guided label propagation and manual refinement. The pelvic and fetal segmentation models included 20 and 10 labels, respectively (Fig. 1). The networks were trained on 57 datasets and evaluated on 10, using a combined Dice and cross-entropy loss function, with increased loss weighting for landmark classes, over 100,000 iterations.

Following segmentation, each predicted landmark was processed by extracting the largest connected component and computing its centroid, which was then used to derive the respective biometry measurements.

The trained model weights, segmentation, and biometry extraction scripts are made publicly available as a standalone Docker application in the auto-proc-svrtk toolbox repository^12^.

### 2.4 Analysis of cephalopelvic measurements in normal term cohort

Subsequently, the trained networks were applied to segment 206 normal control datasets acquired at 0.55T MRI from term pregnancies between 36–40 weeks’ gestation. All predicted landmark segmentations were reviewed and manually refined where necessary by a single observer with more than 5 years of experience in fetal MRI. The resulting biometry values were used to analyse variations in cephalopelvic measurements at term. In addition, we calculated a cephalopelvic index (CPI) to estimate the risk of cephalopelvic disproportion (CPD). While no universally accepted formula for CPI exists, we followed the approach of Korhonen et al. [6], defining CPI as: *CPI* = (*HC − IC*) + (*HC − MC*) where HC is the fetal head circumference, IC is the pelvic inlet circumference, and MC is the midpelvis circumference. Higher CPI values are associated with increased CPD risks due to larger head in comparison to pelvic dimensions.

For all extracted measurements, analysis of variance (ANOVA) was conducted to assess associations with maternal characteristics and delivery-related parameters. At the time of analysis, delivery outcome data were available for 172 cases: 63 normal vaginal deliveries, 45 instrumental vaginal deliveries, 8 elective caesarean sections, and 56 emergency caesarean sections.

## 3 Experiments and Results

### 3.1 Proposed cephalopelvic biometry protocol

The proposed cephalopelvic biometry protocol is presented in Fig. 2 (the network outputs for one of the cases). The biometry was inspected in both 2D T2w planes and 3D model view by clinicians with extensive experience in fetal MRI and confirmed to be acceptable. The pelvis bone and fetal head labels were also confirmed adequate.

**Fig. 2.**
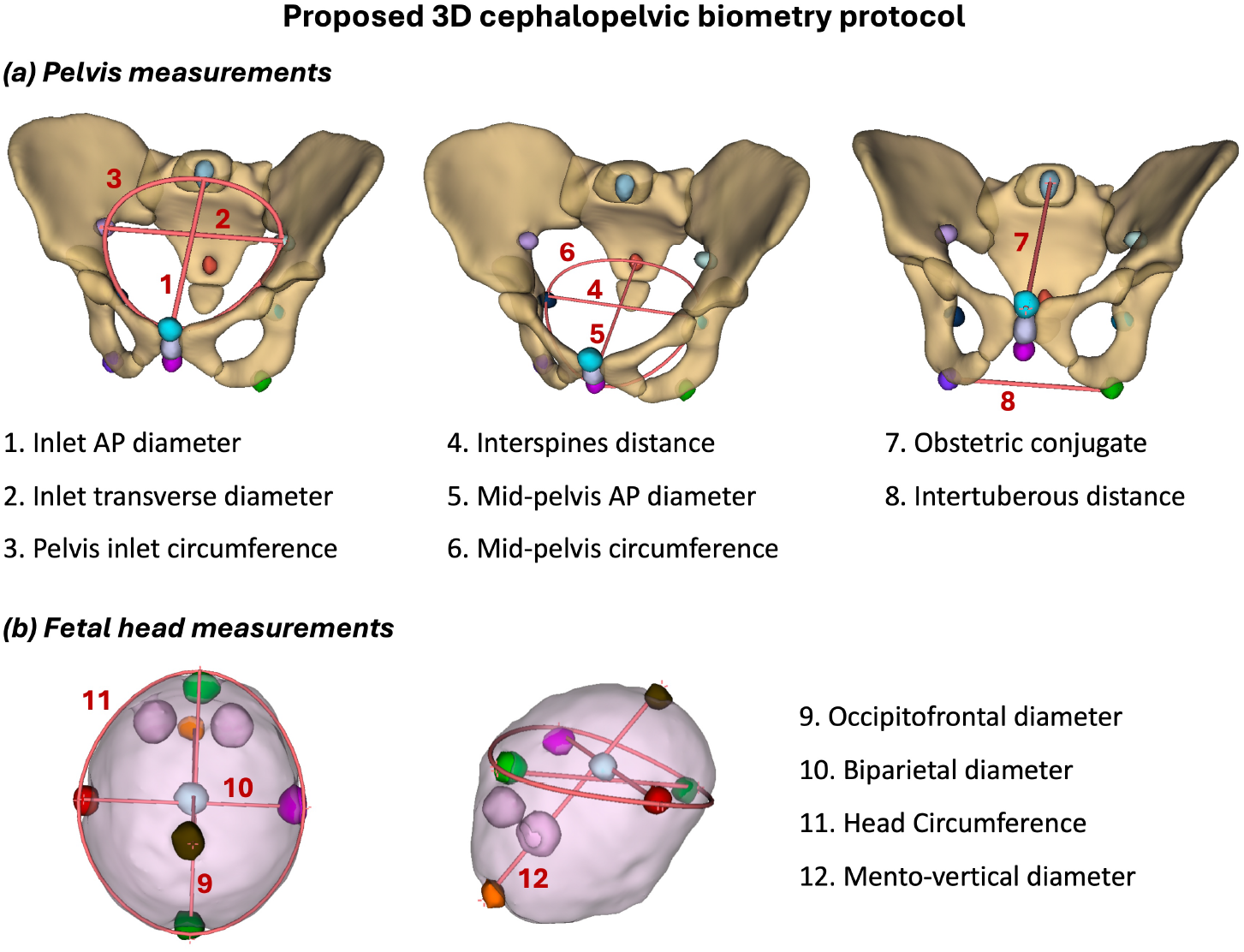
Proposed cephalopelvic biometry protocol for 3D T2w fetal MRI at term: pelvis (a) and fetal head (b) measurements.

### 3.2 Evaluation of automated cephalopelvic biometry pipeline

The results of the quantitative evaluation of the proposed biometry pipeline are summarised in Fig.3, presenting absolute landmark localisation error, intraclass correlation coefficient (ICC), and both absolute and relative errors in the derived biometric measurements across 10 test cases. Overall, the pipeline demonstrated robust performance, with all pelvic and fetal landmarks successfully localised in each test case. Examples of the resulting biometry for two representative cases - illustrating different pelvic morphologies and fetal head positions - are shown in Fig.4.

**Fig. 3.**
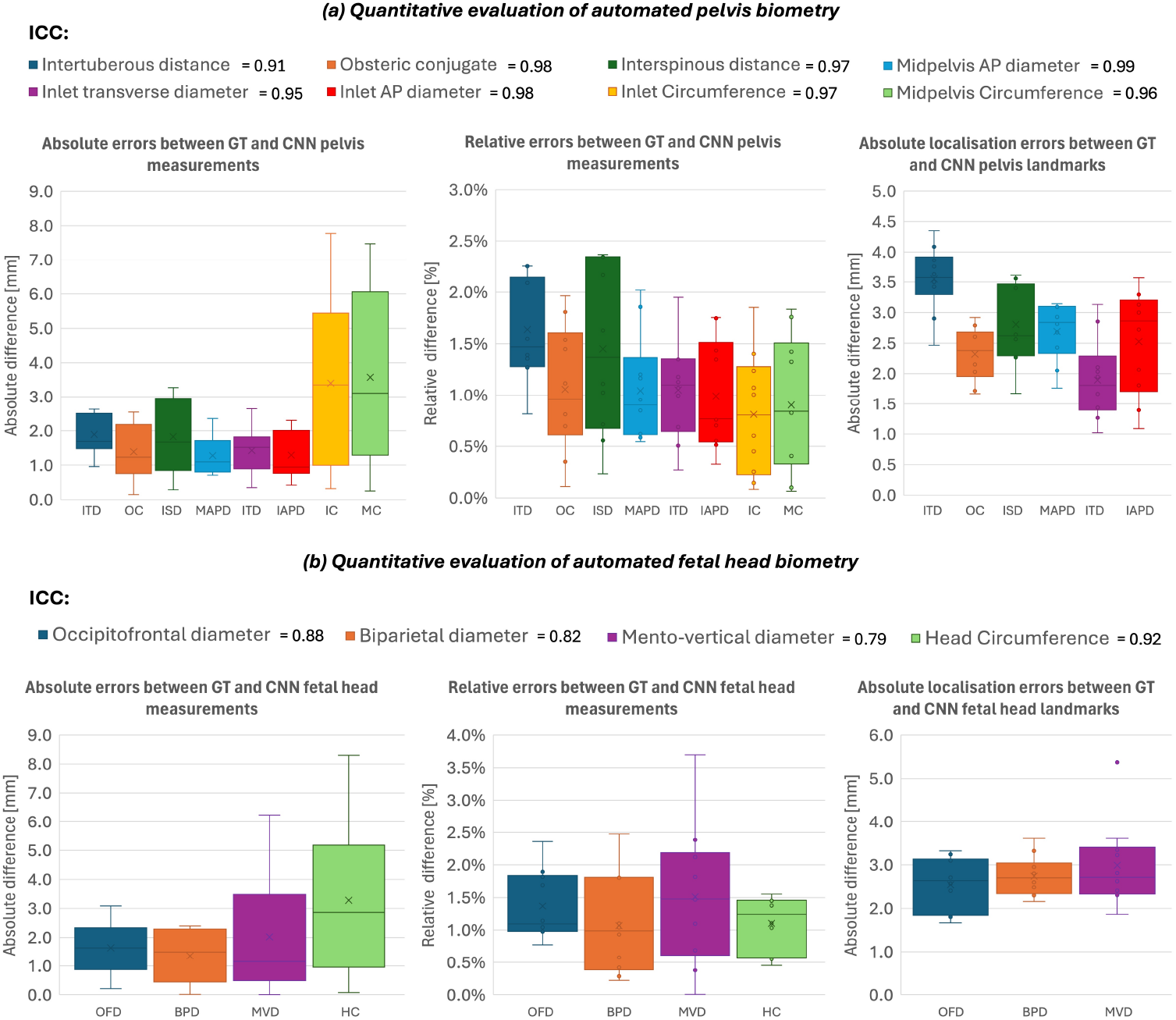
Quantitative evaluation on 10 test cases: comparison of the pelvis (8) and fetal head (4) biometric measurements between the network (CNN) outputs and semimanual ground truth (GT) landmarks. Relative and absolute measurement errors, landmark localisation errors and ICC.

**Fig. 4.**
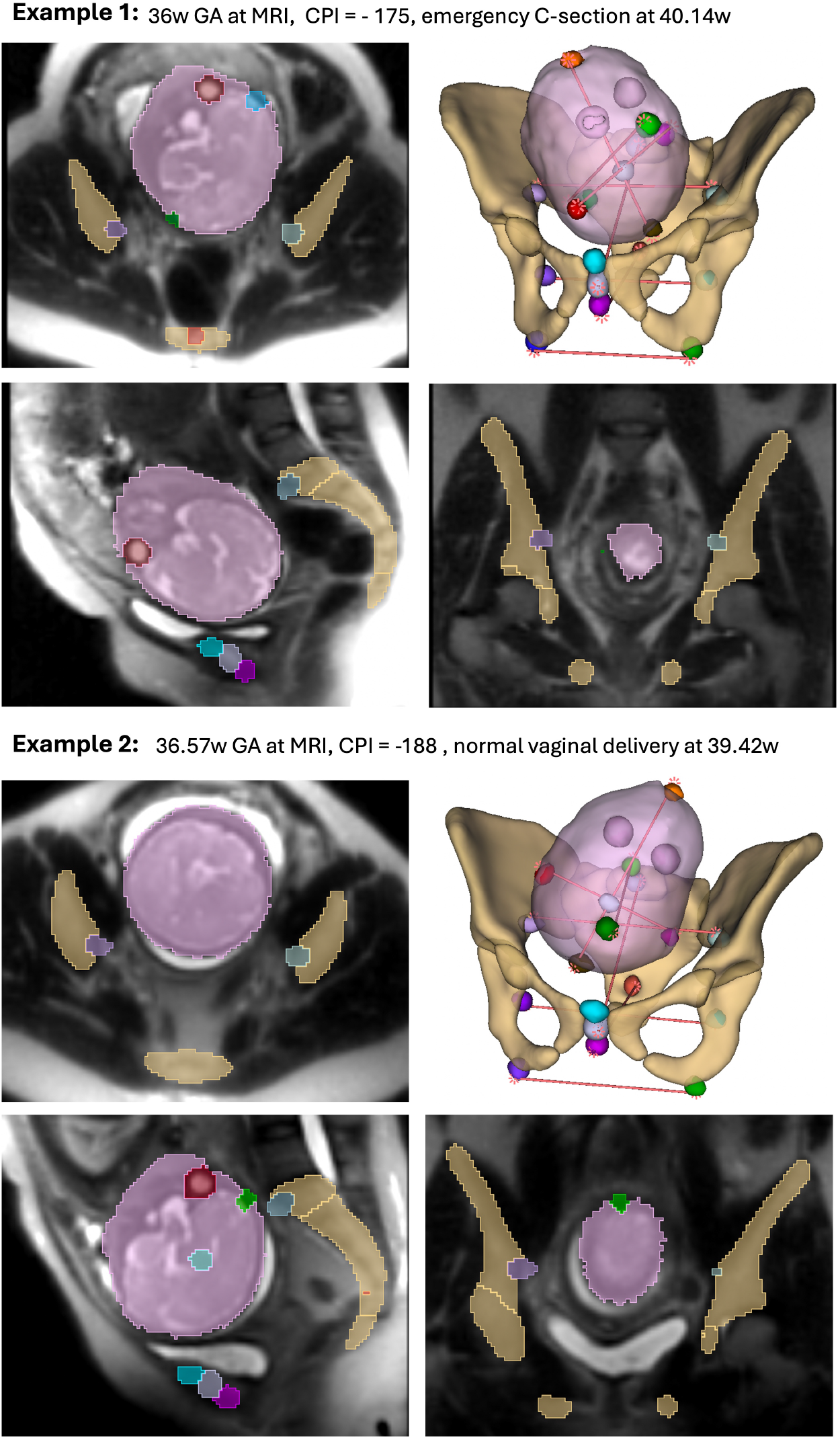
Examples of segmented 3D models of automated biometry for cases with varying pelvis shape and fetal head positions.

The localisation errors relative to the semi-automated ground truth landmarks varied between 2–4 mm for the pelvis and 2–4 3mm for the fetal head. These deviations can be partly attributed to the inherent uncertainty in 3D landmark placement due to the lack of distinct anatomical features—affecting even the ground truth annotations. Another contributing factor is the intrinsically suboptimal image quality at 0.55T, which reduces the visibility of bony structures. The intraclass correlation coefficients with the ground truth measurements were, on average, above 0.8 (corresponding to “excellent” agreement), while relative measurement errors were below 2.5% for pelvic parameters and below 3.0% for fetal head measurements.

These results confirm the feasibility of deep learning-based 3D biometry of the pelvis and fetal head in 0.55T T2-weighted MRI for computing conventional cephalopelvic disproportion parameters. However, the variability in measurements highlights the inherent limitations and oversimplified nature of 2D-based approaches. A more comprehensive assessment using statistical shape analysis may offer improved accuracy and clinical insight.

### 3.3 Analysis of cephalopelvic measurements in normal term cohort

In order to demonstrate the general utility and feasibility of the proposed pipeline we run biometry extraction for 206 term datasets. The graphs in Fig. 5.a illustrate the corresponding examples of the extracted cephalopelvic circumference measurements and the derived cephalopelvic index (CPI). Minor manual refinement of landmarks was necessary in 46 datasets (22%) due to reduced visibility of anatomical structures. As expected, substantial inter-subject variation in pelvic measurements was observed and found to correlate significantly with maternal height (*p <* 0.0001). All fetal head measurements demonstrated a strong correlation with gestational age (*p <* 0.0001). The computed CPI also showed significant associations with both maternal height and gestational age. Notably, there are no established CPI ranges for specific birth outcomes or universally accepted CPI formula.

**Fig. 5.**
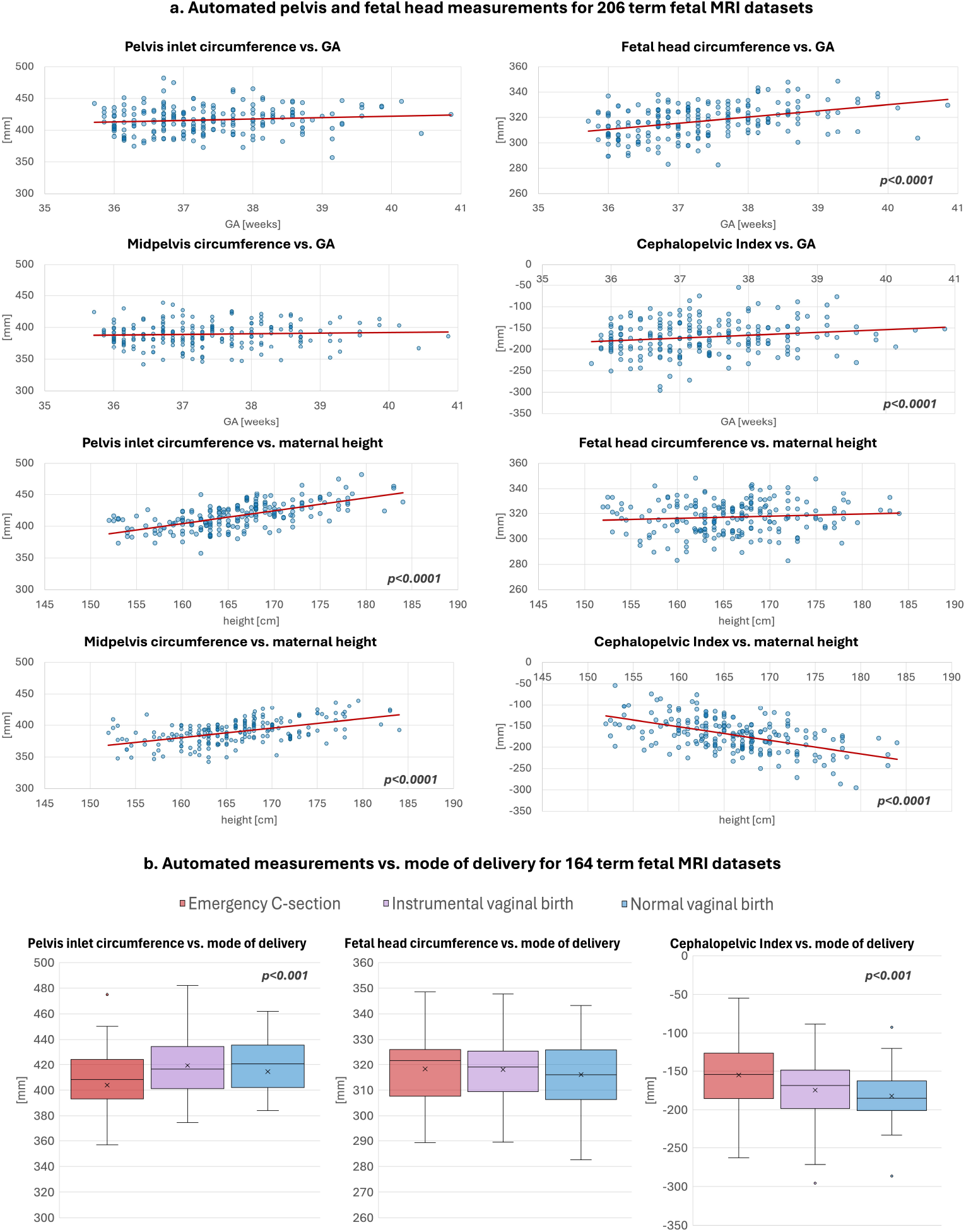
Analysis of automated measurements: maternal pelvis and fetal head biometry and derived cephalopelvic index vs. GA and maternal height for 206 term datasets (a) and comparison vs. mode of delivery for 164 term datasets with known outcomes (red - 56 emergency C-section, lilac - 45 instrumental vaginal delivery, blue - 63 normal vaginal delivery) (b).

Analysis of the 164 datasets with known delivery outcomes (Fig. 5.b) revealed statistically significant associations between pelvic circumference measurements (*p <* 0.001), cephalopelvic index (*p <* 0.001), and mode of delivery. Notably, the emergency caesarean section group exhibited the highest CPI values, followed by the instrumental vaginal delivery group, with the lowest values observed in the normal vaginal delivery group. This trend supports the established understanding that reduced disparity between fetal head and pelvic dimensions is associated with increased risk of cephalopelvic disproportion (CPD) and prolonged labour [11].

These preliminary findings support the utility of the proposed automated biometry pipeline for large-scale fetal MRI studies. They also provide a foundation for comprehensive multi-parametric analyses incorporating additional measurements (e.g., fetal shoulder and abdominal diameters, head engagement), maternal characteristics (e.g., ethnicity, age), and detailed delivery outcomes including fetal weight, maternal and fetal complications, and the duration of labour stages.

## 4 Discussion and Conclusions

This work presents the first prototype methodology for fully automated extraction of maternal pelvis and fetal head biometric measurements from 3D T2-weighted MRI, derived from conventional protocols for assessing the risk of cephalopelvic disproportion at term. The pipeline is based on two 3D UNet networks that segment 12 pelvic and 6 fetal landmarks, which are subsequently used to compute 12 cephalopelvic biometry measurements. In addition, the network segments maternal pelvic bones and the fetal head in order to improve performance and provide anatomical references. The networks were trained on 57 0.55T datasets with semi-manually generated ground-truth labels and quantitatively evaluated on an independent set of 10 test cases. We used the 0.55T datasets because this work is conducted as a part of the MiBirth study and the modern large bore low field scanners are particularly suitable for scanning pregnant women at term [4].

The pipeline demonstrated robust performance for both fetal head and maternal pelvis measurements, achieving ICC above 0.8, absolute errors below 5 mm, and relative errors under 3%. We also performed the general feasiblity analysis based on qualitative evaluation on 206 term datasets. It showed that only minor refinements were required in 22% of cases. The derived cephalopelvic index was significantly correlated with both maternal height and gestational age (*p <* 0.0001). Analysis of measurement associations with delivery mode in a subset of 164 cases with known delivery outcomes further demonstrated that higher CPI values were significantly associated with emergency caesarean sections (*p <* 0.001). Yet, the observed high inter-subject variance in the measurements highlights the need for a more sophisticated multi-factor analysis approach for prediction of delivery outcomes.

Future work will focus on expanding the biometry protocol to include additional pelvic and fetal features, computing relative fetopelvic positional metrics, and performing statistical shape analysis. This will also include assessment of inter- and intra-observer variability in manual ground truth measurements. We aim to integrate automated comparison against normative ranges, perform detailed analysis vs. maternal parameters and generate structured reports highlighting potential patient-specific risk factors. Finally, the pipeline will be applied to 500 term datasets acquired as part of the MiBirth study and incorporated into the birth mode prediction analysis. In order to ensure the added value and utility for wider clinical translation, we will also adapt this work to higher field strength imaging.

## Data Availability

The individual fetal MRI datasets used in this work are not publicly available due to ethics regulations.

## Acknowledgments

We thank everyone who was involved in acquisition and analysis of the datasets and all participating mothers and families.

This work was supported by the MRC grant [MR/X010007/1], the NIHR Advanced Fellowship awarded to Lisa Story [NIHR30166], the Wellcome Trust, Sir Henry Wellcome Fellowship to Jana Hutter [201374/Z/16/Z], DFG Heisenberg funding [502024488], the UKRI, FLF to Jana Hutter [MR/T018119/1], DFG Heisenberg [502024488] the High Tech Agenda Bavaria to Jana Hutter, the Wellcome/ EPSRC Centre for Medical Engineering at King’s College London [WT 203148/Z/16/Z], the NIHR Clinical Research Facility (CRF) at Guy’s and St Thomas’ and by the National Institute for Health Research Biomedical Research Centre based at Guy’s and St Thomas’ NHS Foundation Trust and King’s College London.

The views expressed are those of the authors and not necessarily those of the NHS, the NIHR or the Department of Health.

## Footnotes

9 SVRTK package: https://github.com/SVRTK/SVRTK

10 ITK-SNAP tool: http://www.itksnap.org/

11 3DSlicer tool: https://www.slicer.org/

12 auto-proc-svrtk toolbox: https://github.com/SVRTK/auto-proc-svrtk

## Notes

### Competing Interest Statement

The authors have declared no competing interest.

### Author Declarations

This study employs fetal MRI data with research ethics committee (REC) approval by the Health Research Authority: MiBirth: MRI imaging at term for prediction of the mode of birth study (REC: 23/LO/0685). All experiments were performed in accordance with relevant guidelines and regulations. Informed written consent was obtained from all participants.

